# Rehabilitation in Survivors of COVID-19 (RE2SCUE): a non-randomized, controlled and open study

**DOI:** 10.1101/2022.10.10.22280907

**Authors:** Maria Cristine Campos, Tatyana Nery, Ana Elisa Speck, Maiqueli Arpini, Moisés Moraes Antunes, Ana Cristina de Bem Alves, Naiara de Souza Santos, Maria Paula Pereira Matos, Nelson Schmidt Junior, Letícia Roehe Bicca, Camila Mascarelo Panisson, Mariana Alves Freitas, Fernando Diefenthaeler, Heloyse Uliam Kuriki, Vanessa Damin, Rodrigo Oliveira Da Rosa, Josiane Bueno Gress, Ione Jayce Ceola Schneider, Danielle Soares Rocha Vieira, Livia Arcêncio, Aderbal Silva Aguiar

## Abstract

The sequelae of coronavirus disease-2019 (COVID-19) are another socio-economic problem of the pandemic. Fatigue and dyspnea are the most prevalent symptoms. It is not known whether exercise can be used to treat long COVID-19. This study aimed to investigate the effects of an 8-week face-to-face rehabilitation program on COVID-19 compared to a remote monitoring group. A total of 37 participants (24.3% hospitalized) were assessed before and after rehabilitation (n=22; 40.8±10.0 years) or remote monitoring (n=15; 45.4±10.5 years). The participants were allocated according to their preferences. Both groups showed improved fatigue and exercise capacity (Incremental Shuttle Walk Test). Participants in the face-to-face rehabilitation group showed improved dyspnea (Pulmonary Functional Status and Dyspnea Questionnaire), anxiety (Hospital Anxiety and Depression Scale), attention, and short-term memory (d2-R and Rey’s Auditory-Verbal Learning Test). Of the main sequelae, fatigue improves naturally, whereas dyspnea requires rehabilitation. Our results demonstrated the benefits of exercise for COVID-19 sequelae.

## INTRODUCTION

The severe acute respiratory syndrome coronavirus 2 (SARS-CoV-2) causes coronavirus disease-2019 (COVID-19) and is responsible for developing long-term sequelae^1–3^. Sequelae or persistent symptoms occur even in individuals with mild disease, no symptoms^4^, or vaccinated^5^. “Post-acute COVID-19 syndrome” is referred to for symptoms that persist 4 to 12 weeks after the onset of symptoms, and “post-COVID-19 syndrome” or “Long COVID-19” beyond 12 weeks^6^.

Fatigue and dyspnea are the main sequelae of COVID-19^7–11^. A meta-analysis of studies with hospitalized (n=15.244) and non-hospitalized (n=9.011) COVID-19 survivors concluded that fatigue and dyspnea were the most prevalent symptoms (35– 60%), followed by cough (20–25%)^12^. Fatigue is accompanied by cognitive and neuropsychiatric manifestations, such as brain fog, memory and attention impairment, headache, joint pain, muscle pain, insomnia, anosmia, and dysgeusia^10,13–15^. Anxiety and depression are also common and worsen over time^1,14^.

Impaired lung function accompanied by dyspnea may persist after SARS-CoV-2 infection^16^. Ground-glass opacities on chest computed tomography (CT), fibrotic changes, and reduced carbon monoxide diffusing capacity are found up to 1 year after infection^17^. Peripheral muscle weakness and reduced physical performance have been diagnosed in patients recovering from COVID-19 pneumonia without previous locomotor impairments^18^. Limitation in activities of daily living (ADLs) was observed in up to 45% of the survivors (n=1.142), whose main symptoms were fatigue and dyspnea^19^.

Reduced exercise capacity in up to 3 months has been identified in patients hospitalized for the disease. Of the 200 participants, 49.5% presented with oxygen consumption (VO_2_) below the expected percentage. The reasons for interruption in the cardiopulmonary exercise test (CPET) were lower limb fatigue (93%), dyspnea (5%), and exercise-induced arrhythmia (2%)^20^. Exercise intolerance after COVID-19 appears to improve over time^21^. In contrast, the proportion of participants with reduced performance in the 6-minute walk test (6MWT) was 14% (174/1,254) at 6 months and 12% (147/1,248) at 1 year, demonstrating that in many cases, there is no significant recovery^1^. In addition, reduced VO_2_ was observed almost 1 year after infection, even in mild COVID-19 patients without cardiopulmonary diseases^22^.

Rehabilitation improves symptoms, increases cardiorespiratory fitness, enhances the quality of life, and reduces morbidity and mortality in patients with respiratory diseases^23^. In COVID-19 patients, rehabilitation programs can mitigate the sequelae and burden of the disease^24^. Increased functional capacity, improved lung function, improved quality of life, and reduced anxiety, but not depression, were described after a 6-week rehabilitation program in 36 elderly COVID-19 survivors (69.4 ± 8.0 years) compared to the control group (n=36; 68.9±7.6 years)^25^. The program included respiratory muscle training, diaphragmatic training, coughing exercises, and stretching but did not include aerobic and/or resistance exercises. Another study investigated these components with an 8-week training protocol using aerobic (80% of lactate threshold) and resistance (40% of 1-repetition maximum) exercises. According to the results, benefits for cardiopulmonary and musculoskeletal functions were observed in a sample of 50 participants (55.8 ± 9.7 years) 3 months after hospital discharge^26^. However, there was no control group to compare and establish the treatment efficacy. In another 6-week aerobic and strength exercise rehabilitation study for critically ill COVID-19 survivors reduced dyspnea and increased exercise capacity were observed for the rehabilitation group (n=13; 57.6 ± 10.1 years) and for the control group (n=13; 56.8 ± 8.7 years), which did not exercise^27^.

The effects of rehabilitation in treating patients with persistent symptoms of COVID-19 have not been fully elucidated^24^. In addition, evidence has shown an effect on physical function in elderly^25^ or hospitalized subjects^24,26,27^, with few references to cognitive aspects^28^. In this sense, the present study aimed to evaluate the effects of a face-to-face physical exercise rehabilitation program in patients with persistent symptoms of COVID-19 on fatigue and dyspnea, exercise capacity, pulmonary function, functional status, cognitive function, symptoms of anxiety and depression, and peripheral muscle strength.

## METHODS

### Study design

This was a clinical, non-randomized, controlled, and open study. The protocol followed the guidelines of the Standard Protocol Items for Randomized Trial*s* (SPIRIT), and the results are reported in accordance with the Consolidated Standards of Reporting Trials (CONSORT).

### Participants

Individuals older than 18 years, of both sexes, with a confirmed diagnosis of COVID-19 within 6 months by reverse transcription polymerase chain reaction (RT-PCR) or immunochromatographic test were included after the acute phase of the disease and the presence of at least one of the following symptoms: fatigue, dyspnea, cough, muscle, and/or joint pain.

Individuals with previous respiratory diseases (asthma, chronic obstructive pulmonary disease, and/or pulmonary fibrosis), moderate-to-severe heart disease (New York Heart Association [NYHA] III or IV), neurological, or osteoarticular disease were excluded. Subjects with symptoms of cardiac origin and changes in the resting electrocardiogram with contraindication to physical exertion, reported by a cardiologist, or with cardiovascular disease without regular medical follow-up.

The research was conducted at the Policlínica Regional of the state government in Araranguá, in the extreme south of Santa Catarina, Brazil. Patients were recruited via telephone contact and WhatsApp messages from the COVID-19 registry of the region’s health department. In addition, the survey was disseminated on social media to recruit individuals who were interested in participating in the study.

### Interventions

The face-to-face rehabilitation group participated in treatment supervised by physical therapists for 8 consecutive weeks, twice a week, with an average duration of 80/min/session. Aerobic exercise was performed on a treadmill at a moderate intensity. Participants performed 5 min of warm-up and recovery and 30 min of training at the target intensity. The initial intensity was 75% of the speed achieved in the incremental shuttle walk test (ISWT). This intensity was subsequently adjusted to 60–75% of the reserve heart rate calculated by the Karvonnen method, and the perceived exertion was maintained between 4–6 on the modified BORG scale.

Resistance exercises were performed with an initial intensity of 80% of 10 maximum repetitions (10 RM) and intervals of 1–2 min between 3 sets of 10 repetitions for the trunk and upper and lower limbs. Finally, the trained muscles were stretched (30 s) at the end of the session.

Remote monitoring participants received an original booklet containing general guidelines on physical activity, breathing exercises, energy conservation techniques, respiratory etiquette, nutritional information, and water intake (Supplementary Notes). Every 15 days, contact was made by a video call or WhatsApp message to reinforce the guidelines and monitor health status.

The evaluations were performed in the morning on two different days with an interval of 48 h. After the initial week of assessment, the participants began face-to-face rehabilitation or 8-week remote monitoring. The collection and interventions were conducted from April 2021 to April 2022.

Rehabilitation was carried out in the afternoon, and monitoring of the remote group was carried out at a pre-established time, according to the availability of each participant. In addition, the researchers underwent rigorous training to standardize assessment and intervention measures.

### Primary Outcomes

Fatigue, dyspnea, and exercise capacity were the primary outcome measures. The Modified Pulmonary Functional Status Dyspnea Questionnaire was used to assess dyspnea and fatigue in ADLs^29^. Exercise capacity was evaluated using the ISWT, considering the greatest distance covered after two tests with an interval of 30 min^30,31^.

### Secondary Outcomes

Lung function, functional status, symptoms of anxiety and depression, cognitive function, handgrip strength, and knee extensor strength were secondary outcome measures.

Pulmonary function testing was performed according to international guidelines, fulfilling acceptability and reproducibility criteria^32^. A previously calibrated portable spirometer (Koko Sx 1000 Nspire) was used in this study. Based on the values predicted by Pereira (2007)^33^, data were analyzed on forced vital capacity (FVC), forced expiratory volume in 1 s (FEV_1_), FEV_1_/FVC ratio, and peak expiratory flow (PEF).

Post-COVID-19 Functional Status scale was used to assess the functional status, and the classification was performed considering “0” as no functional limitation up to “4” as the worst level that corresponds to severe functional limitation^34^.

Anxiety and depression symptoms were assessed using the Hospital Anxiety and Depression Scale (HADS). Analyses were performed considering the two domains and the total score of the scale^35^.

Cognitive function was assessed by considering memory and attention, the Brazilian version of the Rey Auditory Verbal Learning Test (RALVT)^36^, and the d2-R^37^. The variables verbal short-term memory (A1), verbal episodic short-term memory (A6), verbal episodic long-term memory (A7), and learning-related memory (REC) were considered for RAVLT analyses. The performance variables in d2-R were obtained using the raw score of concentration performance (CP), number of target objects processed (TOP), and % error (E%), and the respective classification was performed based on the percentile according to sex, age, and education.

Handgrip strength of the dominant limb was measured following the recommendations of the American Society of Hand Therapy using a Jamar® analog dynamometer. Three maneuvers were collected at a 1-minute interval, and the subject’s maximum and predicted values were considered^38^.

Knee extensor strength (kgf) was assessed using maximal voluntary isometric contraction (MVIC) of the dominant lower limb^39,40^. The participants were seated in a chair with a backrest, upper limbs crossed over the chest, 90º of hip flexion, and 60º of knee flexion. The tested lower limb was coupled to a load cell through a shin guard, and the load cell was fixed to the chair by an inextensible chain. The MVIC maneuvers with a 1-minute interval were performed until there was a difference of less than 10% between the two highest measurements.

The electrical activity (in microvolts) of the rectus femoris, vastus lateralis, and vastus medialis was recorded using surface electromyography during MVIC. The electrodes were fixed according to the specifications of the Surface ElectroMyoGraphy for the Non-Invasive Assessment of Muscles. The digital converter used was the New Miotol Miograph software (force load cell model; Miotec Biomedical Equipment, Porto Alegre, Brazil).

### Sample size

The sample calculation was performed according to the results of functional capacity assessed by the 6MWT in the study by Liu et al.^25^. Considering a power of 80%, a significance level of 5%, and a loss of 10%, the estimate was 41 participants per group.

### Allocation

Volunteers chose to participate in the face-to-face rehabilitation or remote monitoring based on their availability and preference.

### Statistical analysis

Descriptive analyses are presented as mean and standard deviation or median and 25-75 quartile, absolute value, and relative frequency. In addition, the delta (Δ) corresponding to the difference between the final and initial moments was reported. The Shapiro-Wilk test was used to test the normality of the distribution of variables. Comparisons between groups were performed using the unpaired t-test or Mann-Whitney U test for numerical variables and chi-squared or Fisher’s exact test for categorical variables. Pre- and post-intragroup results were compared using the paired t-test or Wilcoxon test. Delta (Δ) was considered for the analysis of intergroup outcomes. The effect size was calculated as “small” (0.20−0.50), “medium” (0.50−0.80), or “large” (>0.80)^41^. A significance level of 5% was considered statistically significant. Analyses were performed using the SPSS program, version 22.0, and graphs were generated using GraphPad Prism. version 8.0.2.

### Ethical Aspects

This study was approved by the Research Ethics Committee of the Universidade Federal de Santa Catarina (CAAE:38682820.0.0000.0121) and was registered in the Brazilian Registry of Clinical Trials (RBR-5p8nzk6). The study was conducted in accordance with Resolution 466/2012 of the National Health Council. All the participants provided written informed consent.

## RESULTS

A total of 3,660 individuals diagnosed with COVID-19 were invited to participate in the study through text messages and social media. After telephone screening, 104 eligible participants underwent face-to-face assessments. Of these, four withdrew from participating at the time of the evaluations, and 23 were excluded. A total of 77 articipants were included; 35 withdrew from participating, and five were considered losses (one due to a high-risk pregnancy, one due to a car accident, a new respiratory infection, and two orthopedic injuries not related to the present study), and 37 participants were included (Figure 1). The reasons for dropping out were that the opening hours were incompatible with the participant’s working hours, lack of transport to the rehabilitation service, and clinical improvement. The participants in the face-to-face rehabilitation group performed an average of 13 sessions of the 16 proposals.

**Figure 1.**
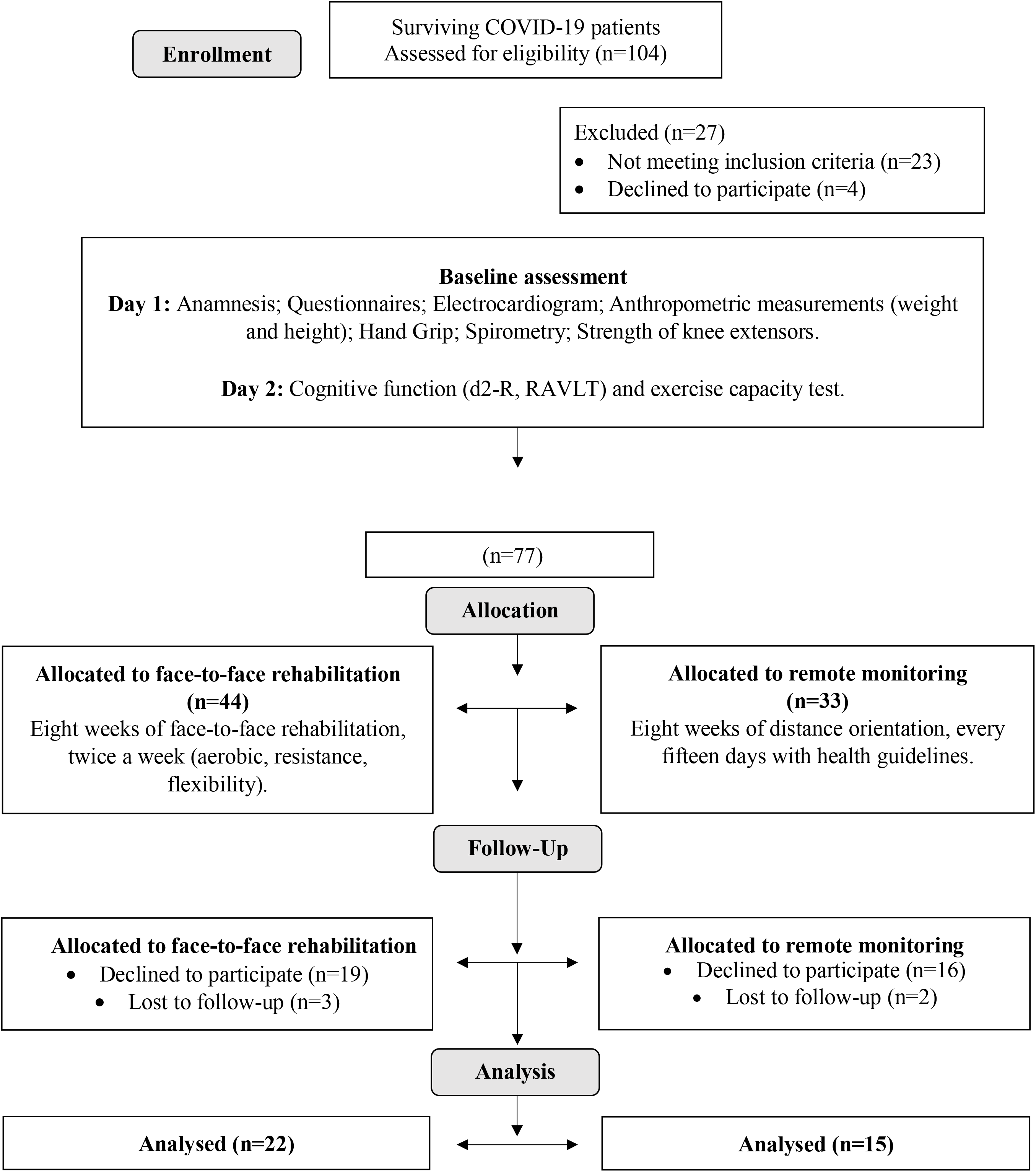
Flowchart of the eligibility, allocation, monitoring, and analysis process of study participants (n=37). Source: by the authors (2022), adapted from CONSORT.

Table 1 presents the characteristics of the 37 participants: 22 were allocated to the face-to-face rehabilitation group and 15 to the remote monitoring group. There were no differences in the sociodemographic characteristics between the groups. In general, the body mass index (BMI) showed overweight. The most prevalent comorbidity was depression, followed by hypercholesterolemia, diabetes mellitus (DM), and systemic arterial hypertension (SAH). Only one participant from each group went to the intensive care unit (ICU), and most were evaluated within 3 months of diagnosis.

**Table 1.** Sociodemographic and anthropometric characteristics of the participants (n=37). Legend: Data expressed as mean and standard deviation (SD) or median [quartile 25-75]. Categorical variables are in absolute value and relative frequency (%). BMI: body mass index. DM: diabetes mellitus. ICU: intensive care unit. p: p value for comparison between groups (Chi-squared or Fisher’s exact test and t-test or Mann-Whitney).

|  | Face-to-face rehabilitation<br>group (n=22) | Remote monitoring<br>group (n=15) | *p value* |
| --- | --- | --- | --- |
| **Age (years)** | 40,8 (10,0) | 45,4 (10,5) | 0,187 |
| **Sex** |  |  | 0,385 |
| Female | 12 (54,5%) | 6 (40%) |  |
| Male | 10 (45,5%) | 9 (60%) |  |
| **BMI (Kg/m<sup>2</sup>)** | 27,2 (4,4) | 27,2 (4,0) | 0,996 |
| **Self-declared skin color** |  |  | 0,481 |
| White | 15 (68,2%) | 12 (80%) |  |
| Brown | 7 (31,8%) | 2 (20%) |  |
| **Schooling** |  |  | 0,098 |
| Primary Education | 7 (31,8%) | 3 (20%) |  |
| Complete High School | 10 (45,5%) | 3 (20%) |  |
| Complete Undergraduate | 5 (22,7%) | 9 (60%) |  |
| **Comorbidity** |  |  |  |
| Depression | 7 (31,8%) | 4 (26,7%) | 1,000 |
| Hypercholesterolemia | 5 (22,7%) | 4 (26,7%) | 1,000 |
| DM | 3 (13,6%) | 2 (13,3%) | 1,000 |
| Hypertension | 2 (9,1%) | 3 (20,0%) | 0,377 |
| **Previous smoking** | 4 (18,2%) | 7 (46,7%) | 0,080 |
| **Hospitalization** | 6 (27,3%) | 3 (20,0%) | 0,711 |
| **Intensive Care Unit** | 1 (4,5%) | 1 (6,7%) | 1,000 |
| **Months after COVID-19** |  |  | 0,677 |
| Three months | 17 (77,3%) | 13 (86,7%) |  |
| Between four and six months | 5 (22,7%) | 2 (13,3%) |  |

### Fatigue, dyspnea, and exercise capacity

Figure 2 shows the results for the primary outcomes. Fatigue was significantly reduced in the face-to-face rehabilitation (*p*=0.0001; d=0.62) and remote monitoring *(p*=0.012; d=0.41) groups. There was no significant difference between the initial fatigue score at baseline (*p*=0.938) and in the deltas between groups (*p*=0.292; d=0.17). Dyspnea in ADLs was significantly reduced only in the face-to-face rehabilitation group (*p*=0.001; d=0.54). There was no difference between the baseline dyspnea score (*p* =0.901) and in the deltas between groups (*p*=0.608; d=0.08). Both had a significant reduction in the total PFSDQ score, which showed no difference between the initial score at baseline (*p*=0.853) and in the deltas between the groups (*p*=0.430; d=0.12).

**Figure 2.**
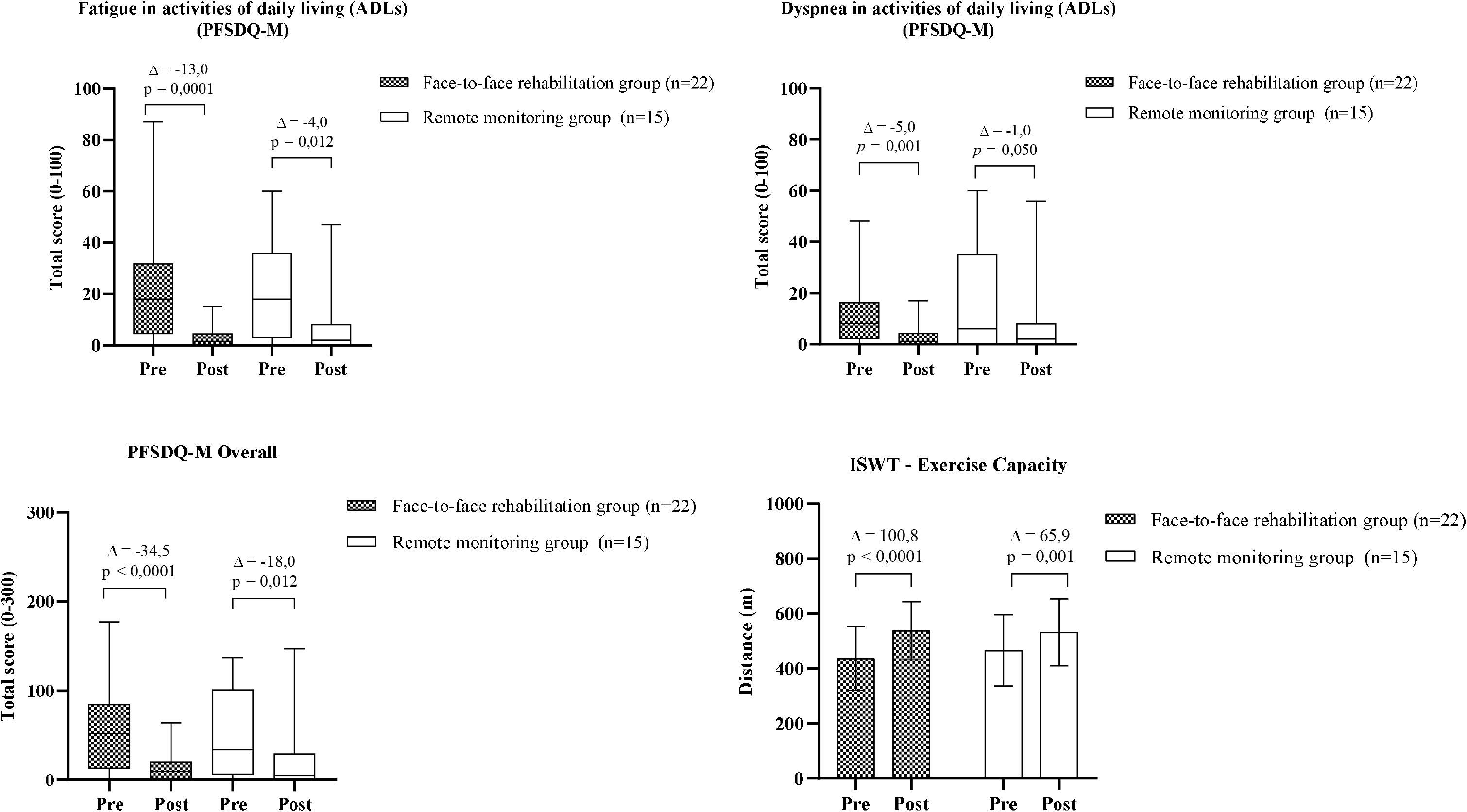
Results of the study’s primary outcomes (n=37). Legend: PFSDQ-M: Pulmonary Functional Status and Dyspnea Questionnaire - Modified version. ADLs: Activities of Daily Living. ISWT: Incremental Shuttle Walk Test. *p*: intragroup p value. Δ: mean (ISWT) and median (PFSDQ-M) of the delta corresponding to the difference between the final and initial moments of the study.

Exercise capacity increased by an average of 100.8 ± 86.1 m in the face-to-face rehabilitation group (*p*< 0.0001) with a large effect size (d=1.17; power 99%). The remote monitoring group had a mean increase of 65.9 ± 61.9 m (*p*=0.001) with a large effect size (d=1.06; power 96%). There was no significant difference between groups for distance covered at baseline (*p*=0.479) or between deltas (p=0.186; d=0.47; power 27%). Among the participants who increased their exercise capacity, the minimum clinical difference of 47.5 m^31^ was observed in 16/22 (72.7%) and 10/15 participants (66.7%) in the face-to-face rehabilitation and remote monitoring groups, respectively, with no significant difference between the groups (*p*=0.728).

### Lung function

Pulmonary function showed a restrictive pattern for five participants (22.7%) in the face-to-face rehabilitation group and one participant (7.7%) in the remote monitoring group, with no significant difference between groups (*p*=0.377). Peak expiratory flow increased significantly (*p*=0.028; d=0.37) only for the face-to-face rehabilitation group (Table 2). Two participants did not complete the pulmonary function test.

**Table 2.** Pulmonary function results (n=35). Legend: Data expressed as median and quartile [25-75]. FVC: Forced Vital Capacity. FEV1: forced expiratory volume in 1 second. PEF: peak expiratory flow. %: percentage of predicted. l: liters. s: seconds. *: p value for intragroup comparison (Wilcoxon). #: p value for intergroup comparison (Man-Whitney). Δ: median (Md) of the delta corresponding to the difference between the final and initial moments. d: effect size for non-parametric data.

| Face-to-face rehabilitation group (n=22) |  |  |  |  |  | Remote monitoring group (n=13) |  |  |  |  |  |  |  |
| --- | --- | --- | --- | --- | --- | --- | --- | --- | --- | --- | --- | --- | --- |
|  | Pre | Post | Md Δ | * *p* | *d* | Pre | Post | Md Δ | * *p* | *d* | # *p*<br>baseline | # *p* Δ | *d* Δ |
| FVC (l) | 3,44<br>[2,99-4,29] | 3,71<br>[3,10-4,41] | 0,10<br>[-0,13-0,21] | 0,194 | 0,22 | 3,91<br>[3,56-4,14] | 4,0<br>[3,55-4,41] | 0,00<br>[-0,05-0,17] | 0,272 | 0,18 | 0,302 | 0,733 | 0,05 |
| FVC<br>(% predicted) | 90,5<br>[79,2-96,7] | 90,0<br>[82,7-99,0] | 2,00<br>[-2,75-5,20] | 0,229 | 0,20 | 93,0<br>[81,5-110,0] | 94,0<br>[85,0-109,5] | 1,00<br>[-1,50-4,00] | 0,228 | 0,20 | 0,151 | 0,797 | 0,04 |
| FEV<sub>1</sub> (l) | 2,92<br>[2,34-3,55] | 3,18<br>[2,41-3,71] | 0,08<br>[-0,07-0,16] | 0,205 | 0,21 | 2,96<br>[2,85-3,39] | 3,00<br>[2,85-3,54] | -0,02<br>[-0,08-0,10] | 0,972 | 0,00 | 0,698 | 0,494 | 0,11 |
| FEV<sub>1</sub><br>(% predicted) | 89,5<br>[82,5-97,7] | 88,5<br>[83,5-99,2] | 1,00<br>[-3,25-4,25] | 0,580 | 0,09 | 94,0<br>[83,0-103,5] | 98,0<br>[85,0-102,0] | 0,00<br>[-2,0-3,00] | 0,782 | 0,04 | 0,559 | 0,864 | 0,02 |
| FEV<sub>1</sub>/ FVC | 84,5<br>[78,0-86,0] | 85,0<br>[77,7-87,0] | 0,00<br>[-1,50-2,00] | 0,887 | 0,02 | 81,0<br>[74,5-83,0] | 79,0<br>[73,5-83,0] | 0,00<br>[-2,00-1,00] | 0,160 | 0,23 | 0,063 | 0,407 | 0,14 |
| PEF (l/s) | 6,98<br>[5,27-8,14] | 7,63<br>[5,76-8,56] | 0,48<br>[-0,34-1,38] | **0,028** | 0,37 | 6,17<br>[5,74-7,90] | 6,25<br>[6,03-7,65] | 0,10<br>[-0,03-0,51] | 0,249 | 0,19 | 0,941 | 0,339 | 0,16 |

### Functional Status

Post-COVID-19 functional status was similar between the groups (Table 3). Therefore, we classified the participants into two categories based on the number of participants. Initially, nine participants (40.9%) from face-to-face rehabilitation presented with mild (7), very mild (1), or no limitation (1), and nine participants (60%) from remote monitoring presented mild (4), very mild (4), or no limitation (1), with no significant difference at baseline between the groups (*p*= 0.325).

**Table 3.** Results of the post-COVID-19 functional status questionnaire (n=37). Legend: Data are expressed as an absolute number corresponding to the number of participants classified dichotomously as “no functional limitation, very mild or mild” or “moderate to severe” limitation. *: p value for intragroup comparison (McNemar test). #: p value for intergroup comparison (Chi-squared test).

|  | Face-to-face rehabilitation group (n=22) |  |  | Remote monitoring group (n=15) |  |  |  |  |
| --- | --- | --- | --- | --- | --- | --- | --- | --- |
|  | Pre | Post | * *p* | Pre | Post | * *p* | # *p*<br>*baseline* | # *p* *post* |
| No functional limitations to Slight | 9 (40,9%) | 13 (59,1%) | 0,219 | 9 (60%) | 10 (66,7%) | 1,000 | 0,325 | 0,738 |
| Moderate to Severe limitations | 13 (59,1%) | 9 (40,9%) |  | 6 (40%) | 5 (33,3%) |  |  |  |

A higher proportion of subjects classified as mild (8), very mild (2), or no limitation (3) was observed after 8 weeks with 13 participants from face-to-face rehabilitation (59.1%; p=0.219) and in the remote monitoring group, 10 participants (66.7%; *p*=1.000) with mild (4), very mild (3) or no limitation (3), with no significant differences between groups (*p*=0.738).

### Anxiety and depression symptoms, assessment of focused attention, memory, and learning

Anxiety was significantly reduced in the face-to-face rehabilitation group (*p*=0.003; d=0.48), which was not observed for the depression outcome (Table 4). In addition, participants who underwent rehabilitation showed a significant improvement in verbal episodic short-term memory (item A6) (*p*=0.039) evaluated by RAVLT, significant improvement in focused attention, verified by the CP index “Concentration Performance” (*p*=0.031) in the d2-R test, and also the TOP index “Processed Target Objects” (*p*=0.031), which represents the execution speed. On the other hand, the participants in the remote monitoring group showed a significant improvement only in the speed of execution, item TOP (*p*=0.016).

**Table 4.** Results of symptoms of anxiety and depression focused attention, memory, and learning (n=37). Legend: Median (Md) and quartile [25-75] data for HADS (Hospital Anxiety and Depression Scale) scores. Numerical data expressed in absolute number and relative frequency (%) correspond to the number of participants classified in typical performance (average or above average) for the tests of focused attention (d2-r), memory, and learning (RAVLT: The Rey Auditory - Verbal Learning Test). TOP Index: target object processed. CP Index: concentration performance index E%: error percentage. A1: verbal short-term memory. A6: verbal episodic short-term memory. A7: verbal episodic long-term memory. REC: recognition (verbal episodic memory related to learning). n = number of participants evaluated. *: p value for intragroup comparison (McNemar test for categorical and Wilcoxon test for numeric). #: p value for intergroup comparison (Chi-squared or Fisher’s exact test for categorical and Mann-Whitney for numeric). Δ: median (Md) of the delta corresponding to the difference between the final and initial moments. d: effect size for non-parametric data.

| Face-to-face rehabilitation group |  |  |  |  |  | Remote monitoring group |  |  |  |  |  |  |  |  |
| --- | --- | --- | --- | --- | --- | --- | --- | --- | --- | --- | --- | --- | --- | --- |
|  | Pre | Post |  |  |  |  | Pre | Post |  |  |  |  |  |  |
| HADS | Md [25-75] | Md [25-75] | *p | d | Md Δ | HADS | Md [25-75] | Md [25-75] | *p | d | Md Δ | # p baseline | # p Δ | d Δ |
| Anxiety<br>(n=22) | 8,5<br>[7,0-11,0] | 5,5<br>[4,0-9,0] | **0,003** | 0,48 | -2,00<br>[-4,24-0,00] | Anxiety<br>(n=15) | 6,0<br>[4,0-10,0] | 7,0<br>[4,0-9,0] | 0,549 | 0,09 | -1,00<br>[-3,00-2,00] | 0,143 | 0,077 | 0,29 |
| Depression<br>(n=22) | 6,5<br>[3,7-9,5] | 7,5<br>[2,7-10,2] | 0,745 | 0,05 | 0,00<br>[-2,25-1,50] | Depression<br>(n=15) | 6,0<br>[4,0-9,0] | 6,0<br>[3,0-7,0] | 0,329 | 0,16 | -2,00<br>[-3,00-2,00] | 0,641 | 0,555 | 0,09 |
| Total score<br>(n=22) | 14,0<br>[10,0-20,7] | 14,5<br>[7,0-17,0] | 0,053 | 0,31 | -2,50<br>[-6,25-1,00] | Total score<br>(n=15) | 14,0<br>[8,0-18,0] | 13,0<br>[7,0-17,0] | 0,462 | 0,12 | -1,00<br>[-5,00-0,00] | 0,328 | 0,577 | 0,09 |
| d2-R test | n (%) | n (%) | *p |  |  | d2-R test | n (%) | n (%) | *p |  |  | # p baseline | # p post |  |
| TOP<br>(n=22) | 13 (59,1%) | 19 (86,4%) | **0,031** |  |  | TOP<br>(n=15) | 6 (40,0%) | 13 (86,7%) | **0,016** |  |  | 0,325 | 1,000 |  |
| CP<br>(n=22) | 14 (63,6%) | 20 (90,9%) | **0,031** |  |  | CP<br>(n=15) | 7 (46,7%) | 12 (80,0%) | 0,063 |  |  | 0,336 | 0,377 |  |
| E%<br>(n=22) | 15 (68,2%) | 19 (86,4%) | 0,125 |  |  | E%<br>(n=15) | 11 (73,3%) | 12 (80,0%) | 1,000 |  |  | 1,000 | 0,670 |  |
| RAVLT test | n (%) | n (%) | *p |  |  | RAVLT test | n (%) | n (%) | *p |  |  | # p baseline | # p post |  |
| A1<br>(n=22) | 16 (72,7%) | 21 (95,5%) | 0,125 |  |  | A1<br>(n=15) | 9 (60,0%) | 11 (73,3%) | 0,687 |  |  | 0,488 | 0,136 |  |
| A6<br>(n=22) | 10 (45,5%) | 17 (77,3%) | **0,039** |  |  | A6<br>(n=15) | 6 (40,0%) | 10 (66,7%) | 0,125 |  |  | 1,000 | 0,708 |  |
| A7<br>(n=22) | 11 (50,0%) | 15 (68,2%) | 0,125 |  |  | A7<br>(n=15) | 8 (53,3%) | 9 (60,0%) | 1,000 |  |  | 1,000 | 0,730 |  |
| REC<br>(n=21) | 13 (59,1%) | 15 (68,2%) | 0,687 |  |  | REC<br>(n=14) | 11 (73,3%) | 7 (46,7%) | 0,375 |  |  | 0,491 | 0,288 |  |

### Peripheral muscle strength

In general, peripheral muscle strength at baseline assessed using a hand grip was above 80% of the predicted value for both groups, with no significant differences (Table 5). There was no significant increase in handgrip strength after 8 weeks of rehabilitation or in the remote monitoring group. In contrast, quadriceps peripheral muscle strength increased significantly (*p=*0.026; d=0.43) only in the remote monitoring group. This increase was accompanied by greater electrical activity in the vastus medialis (*p=*0.008, d=0.52).

**Table 5.** Result of peripheral muscle strength (n=37). Legend: Data expressed as median and quartile [25-75]. MVIC: maximum voluntary isometric contraction. kgf: kilogram force. µV: microvolts. *: p value for intragroup comparison (Wilcoxon). #: p value for intergroup comparison (Mann-Whitney). Δ: median (Md) of the delta corresponding to the difference between the final and initial moments. d: effect size for non-parametric data.

| Face-to-face rehabilitation group |  |  |  |  |  | Remote monitoring group |  |  |  |  |  |  |  |  |
| --- | --- | --- | --- | --- | --- | --- | --- | --- | --- | --- | --- | --- | --- | --- |
|  | Pré | Pós | Md Δ | * *p* | *d* |  | Pré | Pós | Md Δ | * *p* | *d* | # *p*<br>*baseline* | # *p* Δ | *d* Δ |
| Hand Grip (n=22) | 31,0<br>[22,5-46,0] | 31,5<br>[24,0-44,2] | 1,50 | 0,312 | 0,16 | Hand Grip (n=15) | 32,0<br>[29,0-43,0] | 38,0<br>[32,0-41,0] | 2,00 | 0,230 | 0,19 | 0,483 | 0,721 | 0,05 |
| Hand Grip % predicted (n=22) | 90,5<br>[74,7-105,0] | 98,5<br>[86,5-104,0] | 2,00 | 0,322 | 0,16 | Hand Grip % predicted (n=15) | 104,0<br>[80,0-108,0] | 100,0<br>[83,0-113,0] | 7,00 | 0,185 | 0,21 | 0,322 | 0,745 | 0,05 |
| MVIC (kgf) (n=15) | 23,0<br>[14,6-30,5] | 21,7<br>[17,7-31,3] | 2,77 | 0,733 | 0,06 | MVIC (kgf) (n=11) | 25,0<br>[16,4-26,0] | 30,8<br>[19,5-37,2] | 4,81 | **0,026** | 0,43 | 0,775 | 0,223 | 0,09 |
| Rectus Femoris (μV) (n=15) | 115,0<br>[76,4-185,2] | 119,5<br>[89,9-173,5] | 7,81 | 0,394 | 0,16 | Rectus Femoris (μV) (n=11) | 110,6<br>[80,8-212,9] | 163,6<br>[127,0-187,8] | 41,3 | 0,155 | 0,28 | 0,979 | 0,392 | 0,05 |
| Vastus Lateralis (μV) (n=15) | 141,1<br>[74,5-246,0] | 116,8<br>[90,2-245,7] | 24,6 | 0,820 | 0,04 | Vastus Lateralis (μV) (n=11) | 193,7<br>[116,4-295,9] | 205,7<br>[131,9-292,6] | 61,6 | 0,286 | 0,21 | 0,392 | 0,517 | 0,02 |
| Vastus Medialis (μV) (n=15) | 149,0<br>[88,7-218,2] | 115,5<br>[72,1-204,3] | -13,8 | 0,910 | 0,02 | Vastus Medialis (μV) (n=11) | 111,1<br>[66,5-162,5] | 178,1<br>[132,4-232,2] | 54,6 | **0,008** | 0,52 | 0,364 | 0,040 | 0,11 |

## DISCUSSION

In this study, we demonstrated that subjects with persistent symptoms of COVID-19 who participated in an 8-week face-to-face rehabilitation program had reduced fatigue, dyspnea, anxiety, increased exercise capacity, and improved episodic verbal short-term memory and focused attention. In addition, remote monitoring of participants reduced fatigue and increased exercise capacity and quadriceps strength.

To our knowledge, this is the first clinical study of patients with COVID-19 sequelae followed for 8 weeks to investigate the effects of physical exercise on physical and cognitive performance in this population. A recent systematic review^24^ showed that nine observational studies (n=957) and 14 intervention studies (n=469) on rehabilitation in subjects with COVID-19 had been published. In most studies, rehabilitation was carried out during hospitalization and the acute phase of the disease (within the first 4 weeks after the onset of symptoms), or in the post-acute phase (after 4 weeks of the onset of symptoms) lasting 1^42^, 4^43^, and 6 weeks^25^.

### Fatigue and Dyspnea

Fatigue and dyspnea are the most prevalent symptoms in individuals with long COVID-19, hospitalized or not^12^. Fatigue is a common condition after viral infections since the Spanish flu was caused by the influenza virus (H1N1) in 1918^44^, and more recently in survivors of severe acute respiratory syndrome (SARS)^45^ and the Middle East respiratory syndrome (MERS)^46^ caused by coronaviruses. Fatigue is debilitating and can be physical or mental, and is characterized by weakness, lack of energy, impaired concentration, sluggishness, and drowsiness^47^. Despite being common in neurological disorders^48^ and chronic respiratory diseases^49^, its definition and pathophysiology are not fully established^48^. In COVID-19, central, psychological, and peripheral factors have been suggested to be involved in the pathophysiology of fatigue^50^. Due to the multifactorial aspect and impact on ADLs^51^, we used the PFSDQ-M^29^ to assess this outcome in our study.

Fatigue in ADLs had a higher score on the PFSDQ-M when compared to dyspnea and was reduced in both groups. Similarly, a reduction in fatigue with a drop in proportion from 52% to 20% was observed in a cohort of 1,276 COVID-19 survivors aged between 6 and 12 months^1^. However, there is evidence that the proportion of subjects with fatigue 12 weeks or more after diagnosis is 0.32 (95% CI, 0.27, 0.37; p<0.001; n=25,268; I^2^ =99.1%)^52^ and persisting for up to 1 year (33% of 192 survivors)^53^. These data allowed us to assume that, regardless of clinical improvement, a portion of those infected still maintained fatigue symptoms, as was observed in our sample.

Regarding rehabilitation, the reduction of fatigue, evaluated by FACIT-Functional Assessment of Chronic Illness Therapy (improvement of 5±7 points), was described in an observational study with 30 patients undergoing treatment for 6 weeks who performed aerobic and resistance exercises^28^. “Reported” fatigue and dyspnea also reduced after 5-week multidisciplinary rehabilitation with moderate to high-intensity aerobic and resistance exercise in 95 participants^54^. These data, together with our results, suggest that time and exercise appear to be factors that affect post-COVID-19 fatigue.

In our study, dyspnea was significantly reduced only in the face-to-face rehabilitation group. Khurana et al.^27^ also observed symptom improvement for rehabilitation participants, with a mean reduction of −2.0±0.6 points on the Medical Research Council Modified Dyspnea Scale (mMRC), significantly greater (*p*=0.01) than −1.3±0.8 points in the control group. These findings demonstrate the potential effect of rehabilitation on dyspnea caused by COVID-19, which appears to improve with exercise. Above all, a reduction in dyspnea was also observed in the control group in the aforementioned study. Time appears to influence dyspnea improvement. Resolution of the pulmonary inflammatory process may be one of the main reasons for this. A reduction in dyspnea and improved structural changes on chest tomography were observed in a 3-month follow-up of COVID-19 survivors (n=145; 75% hospitalized)^16^. In contrast, another study showed that dyspnea was prevalent during activities (55%) and at rest (23.5%) 7 months after hospital discharge in a cohort of 1,142 survivors^19^. Furthermore, in the cohort by Huang et al.^1^, dyspnea assessed by mMRC increased from 26% (313/1,185) at 6 months to 30% (380/1,271) at 12 months (*p*=0.014). These results corroborate the findings of up to 1 year in CT of the chest and lung function of COVID-19 survivors with fibrotic changes that showed little improvement and decline in diffusion capacity^17^.

Dyspnea or respiratory distress occurs due to several clinical conditions, but it is also a reflection of reduced cardiovascular fitness^55^. In COVID-19, it is related to inflammation of the pulmonary alveoli, thrombosis, microclots, and neuroinvasion^56^. In fact, it was shown that although 86% of 1,099 patients with COVID-19 had chest tomography abnormalities and a low relationship between the partial pressure of oxygen and fraction of inspired oxygen (PaO_2_/FiO_2_), dyspnea was reported in only 18.6%^57^ The variation in the perception of dyspnea may be caused by the neuroinvasive potential of SARS-CoV-2 due to the involvement of the cardiorespiratory center^58^ or higher centers^56^. These data suggest that impaired lung function is not the only mechanism responsible for triggering dyspnea in patients with COVID-19.

An increase in peak expiratory flow was observed after 8 weeks of face-to-face rehabilitation. Tang et al.^43^ observed a significant reduction in dyspnea and an increase in peak inspiratory flow after 4 weeks of “Liuzijue” exercises with breathing exercises associated with body movements in a sample of 33 participants with mild to critical illness. Liu et al.^25^ demonstrated the effects of a 6-week respiratory rehabilitation program. The authors did not investigate dyspnea. Overall, improvements in the lung function were observed. Together with our results, these findings demonstrate that dyspnea is a complex and multifactorial sequela^55^. Considering that exercise is the component responsible for improving dyspnea in patients with respiratory diseases^55^, rehabilitation seems to be necessary to treat this post-COVID-19 symptom.

### Exercise capacity

Both groups showed an increased exercise capacity. Our results were similar to those observed by Daynes et al.^28^ with 30 participants (58±16 years), 87% of whom were hospitalized. Participants in the aforementioned study had a mean initial ISWT distance of 300±198 m and a mean gain of 112±105 m (*p*<0.01) after rehabilitation. In our study, the face-to-face rehabilitation group had better exercise capacity at baseline. Our sample consisted of a smaller proportion of hospitalized subjects. In addition, our participants were relatively younger, and most were evaluated within 90 days of being diagnosed with COVID-19, whereas in another study, the median time was 125 days. These characteristics justify the better exercise capacity of our participants at baseline.

In the study by Khurana et al.^27^, the 6-week rehabilitation was started between the first and fourth week of ICU discharge, with aerobic exercises at 70% of the speed achieved in the 6MWT, resistance exercises for the quadriceps and upper limbs, and breathing exercises for lung expansion. The 14 participants in the intervention group (57.6±10.1 years) were compared with 13 in the control group (56.8±8.7 years). The intervention group, with a baseline of 218.2±52.9 m in the ISWT, had a mean increase of 152.5 m after rehabilitation, which was statistically higher (*p*=0.02) than the control group, with a mean increase of 64.9 m and baseline of 243.6±107.2 m. These results are similar to those observed in the remote monitoring group in our study. This evidence highlights three important issues. First, rehabilitation improves the exercise capacity of subjects affected by COVID-19. Second, patients with severe disease and older age present greater impairment in exercise capacity, and finally, exercise capacity seems to improve for subjects who do not undergo physical rehabilitation but to a lesser extent.

### Functional status

Functional status limitation was observed even after 8 weeks in both groups, and there was no significant improvement after rehabilitation. These findings can be explained by participants’ baseline characteristics. Our sample had a relatively low proportion of hospitalized patients who required intensive care. Above all, it is known that approximately 80% of cases present with the mild disease without the need for hospitalization^59^, which justifies the inclusion of individuals with fewer limitations.

Regardless of hospitalization, COVID-19 affects the functionality of infected individuals. A population-based study showed that middle-aged and elderly patients in the community with mild or moderate COVID-19 were associated with worsening mobility and function^60^. In the study by Nopp et al.^61^, after 6 weeks of rehabilitation, the Post-COVID-19 Functional Status Scale (PCFS) score showed a significant reduction in the median score from 2 (interquartile range, 2–3) to 1 (interquartile range, 0–2)(*p*<0.001) in a sample of 58 patients (46.8±12.6 years), 62% were not hospitalized, with mild to moderate disease.

Our objective was to verify the impact of rehabilitation on the improvement of functional status. We did not investigate which constructs were more limited, such as basic and instrumental activities or social participation and leisure, but we understand that these are important aspects for further research. At least one functional limitation in ADLs was observed in 45% of a cohort of 1,124 participants who were hospitalized for COVID-19^19^, with a greater limitation for occupational activities (22.5%), female sex (odds ratio 1.75, 95%CI:1.38–2.22, *p*<0.001), and ICU patients (odds ratio 2.16, 95%CI:1.34–3.48, *p*=0.001). In addition, those with the highest levels of fatigue and dyspnea also had the most limitations (*r* =0.359–0.684, all *p*<0.001).

### Anxiety and depression

Anxiety was only reduced in the face-to-face rehabilitation group. This result corroborates the findings of Liu et al.^25^, which showed a significant reduction in the anxiety score, but not in depression, for participants in the intervention group. In contrast, the 6-week rehabilitation in the study by Khurana et al.^27^ did not interfere with symptoms of anxiety and depression. In a study by Tang et al.^43^, anxiety and depression were significantly reduced after 4-week rehabilitation with “Liuzijue” exercises.

Anxiety and depression are common manifestations of COVID-19 and have increased significantly over time^1,14,15^. Direct effects of infection with exacerbated immune response and neuroinflammation are described as biological mechanisms, in addition to psychological mechanisms such as social isolation, trauma during infection, incomplete recovery of health, persistent fatigue, and unemployment^1^. The medium-to-long-term increase in frequency in the study by Premarj et al.^14^ suggests that these symptoms are not only persistent after infection but also develop over time. These findings suggest that rehabilitation in patients with COVID-19 may reduce anxiety. However, it does not seem to influence the improvement of depressive symptoms, demonstrating the need for a psychological and psychiatric approach to managing depression in COVID-19.

### Focused attention, memory, and learning

We observed an improvement in the cognitive function of the participants in face-to-face rehabilitation. Focused attention and verbal episodic short-term memory improved after the treatment. In addition, an increase in the TOP index of the d2-R test was observed in both groups. The TOP index represents the speed of work. Therefore, an increase in the TOP index alone, without an increase in the CP index, does not represent an improvement in focused attention but an increase in the speed of test execution.

We evaluated these constructs considering that memory impairment and attention deficits are common manifestations of post-COVID-19 cognitive function^10,15^. According to Premarj et al.^14^, hippocampal and cortical atrophy, hypoxic-ischemic changes, and small vessel disease secondary to inflammation and oxidative stress during COVID-19 could be responsible for cognitive dysfunction. Determining the underlying mechanisms of physical exercise to improve cognitive function is far from one of our goals. However, it is known that moderate-intensity aerobic and/or resistance exercise can improve cognitive function in adults, regardless of baseline cognitive status^62^.

Regarding rehabilitation, only the study by Dayens et al.^28^ investigated the effects on cognition using the Montreal Cognitive Assessment (MoCA). After rehabilitation, the results showed a two-point improvement in the total MoCA score (25±3 to 27±3; *p*<0.01). Although the remote monitoring group in our study showed improvement in the TOP index, which can be explained by the smaller number of subjects in the group, an increase in the prevalence of neurological disorders occurs in post-COVID-19 patients in need of treatment^14^. These findings allow us to conclude that exercise improves the cognitive function of patients with COVID-19 and that it is necessary to manage these manifestations.

### Peripheral muscle strength

Handgrip strength is widely used in rehabilitation as a basic measure of musculoskeletal function. The measurement represents upper limb strength and is also an indicator of global strength^38^. We observed that the initial handgrip strength did not show significant impairment, which justifies an “absence” of improvement. This result is relevant and is represented by the profile of our relatively young sample with a lower hospitalization rate. In the study by Tuzun et al.^63^, lower handgrip strength was observed in patients with severe disease (n=22; 18.26 kgf) than in non-severe cases (n=51; 23.37 kgf) for females. The results demonstrated that disease severity affects handgrip strength in a sex-dependent manner.

Quadriceps strength assessed by MVIC, although widely used for patients with respiratory disease^39^, has a heterogeneous methodology, which limits the comparison of results. Paneroni et al.^18^ evaluated maximal quadriceps strength of the dominant lower limb of COVID-19 patients (n=41; 67.1±11.6 years) at the time of hospital discharge. Quadriceps weakness (<80% predicted) was observed in 86% of the patients, with a mean value of 18.9 kgf. In our study, the baseline quadriceps strength was higher for participants in both groups. Overall, a significant increase in strength accompanied by an increase in electrical activity was observed only in subjects in the remote monitoring group. Considering the similar handgrip strength between the groups, we understand that this finding is not clinically relevant to the functionality of the subjects. In contrast, this finding suggests aspects of SARS-CoV-2 viral pathogenesis in muscle function. Curiously, lower limb fatigue is a limiting factor for exercise in these patients^20^.

Tanriverdi et al.^64^ evaluated the handgrip and knee extensor strength of 48 participants (39.2±7.9 years) after 12 weeks of COVID-19 diagnosis with mild to moderate disease. The handgrip and quadriceps muscle weakness (<80% predicted) were observed in 39.6% and 35.4% of the participants, respectively. In addition, after 3 months of infection, individuals with the moderate disease had lower strength than those with mild disease. In the study by Barbara et al.^26^, 8-week rehabilitation promoted a significant increase in muscle strength, evaluated through one repetition maximum (1 RM) for knee extensors (22.4±10 kgf to 28.3±11 kgf; *p*=0.009) in a sample of 50 subjects (55.8 ± 9.7 years) who started treatment 3 months after hospital discharge.

These findings demonstrate that peripheral muscle strength in COVID-19 may be impaired depending on the severity of the disease. However, the diversity of methodologies and instruments for assessing quadriceps strength limits the comparison between studies and conclusions of this assessment. In addition, the mechanism related to lower limb weakness and fatigue in COVID-19 needs to be investigated in future studies.

### Limitations

Our study had some limitations. First, this was a non-randomized study. In our pilot study, during the non-vaccine phase of the pandemic, we observed low adherence, and for this reason, we offered participants the power of choice to participate in face-to-face rehabilitation or remote monitoring according to their preferences and possibilities during the lockdown period. We understand that this form of allocation interferes with the validity of our results. In contrast, our sample showed homogeneous baseline characteristics. In view of the pandemic moment we are going through, this was a pragmatic strategy^65^ thinking about patient care and adherence. For ethical reasons, the remote monitoring group received general guidelines for practicing physical activity and breathing exercises and therefore received some assistance, which could even justify the improvements observed in this group.

Second, our sample size was relatively small, which may have affected the power of intra- and intergroup comparisons. Many dropouts occurred because the participant’s work schedule was incompatible with the rehabilitation schedule, while others had no means of transport to the rehabilitation site. Others reported clinical improvement and expressed interest in not continuing the research. In fact, non-adherence is a multidimensional process that is difficult to resolve and includes internal factors such as depression and belief in the importance of activities and external factors such as social support and transportation^66^. In addition, health limitations, lack of social support, lack of motivation, and financial difficulties are barriers that impact adherence to pulmonary rehabilitation programs^67^. Finally, technological support is essential for implementing monitoring and remote monitoring systems.

### Clinical implications

In this clinical study, a rehabilitation protocol for treating subjects with sequelae of COVID-19 was proposed, with moderate-intensity aerobic and resistance exercises. Rehabilitation reduced the symptoms and improved exercise capacity and cognitive function. In addition, the protocol has been shown to be safe, feasible, and accessible to scientific and clinical communities. An educational guidebook for the benefit of subjects with persistent symptoms is also available and can be recommended as a complementary educational component of rehabilitation programs.

## Conclusion

An 8-week face-to-face rehabilitation program can reduce symptoms of dyspnea, fatigue, and anxiety, increase exercise capacity, and improve memory and attention in patients with persistent symptoms of COVID-19. In addition, a portion of those infected may show a reduction in symptoms and an increase in exercise capacity over time. However, further studies are required to investigate the factors related to these findings.

## Data Availability

All data produced are available online at https://doi.org/10.17605/OSF.IO/3J7PM

## Acknowledgments

We thank the funding sources, study participants, research and data collection team members, research laboratories involved, and Maria Schmitt Institute (IMAS) of Araranguá.

## Author Contributions

All authors contributed to writing and editing the manuscript. MCC, DSRV, IJCS, LA, and ASAJ made substantial contributions to the conception and design of the study and critically revised the manuscript. AES, TN, MA, MMA, ACBA, NSJ, LRB, CMP, MAF, ROR, NSS, and JBG made substantial contributions to the acquisition of data for the study. FD, MPPM, HUK, and VD made substantial contributions to the interpretation of data. All authors have read and approved the final manuscript.

## Data availability

The datasets of the current study are available in the Open Science Framework (OSF): https://doi.org/10.17605/OSF.IO/3J7PM

## Registration and Protocol

This research was registered in The Brazilian Clinical Trials Registry (ReBEC) under number RBR-5p8nzk6. The clinical protocol is in the PREPRINT format: https://doi.org/10.1101/2021.09.06.21262986

## Competing interests

The authors declare no competing interests.

## Funding

The funding sources were Fundação de Amparo à Pesquisa e Inovação do Estado de Santa Catarina (FAPESC), the National Council for Scientific and Technological Development (CNPq), the Coordination for the Improvement of Higher Education Personnel (CAPES), and the Ministry of Health (Brazil). Funding sources will have no role in the design, conduct, or reporting of this study.

## Notes

### Competing Interest Statement

The authors have declared no competing interest.

### Clinical Trial

RBR-5p8nzk6

### Clinical Protocols

https://www.medrxiv.org/content/10.1101/2021.09.06.21262986v1.full

### Funding Statement

This study was funded by Fundacao de Amparo a Pesquisa e Inovacao do Estado de Santa Catarina (FAPESC), the National Council for Scientific and Technological Development (CNPq), the Coordination for the Improvement of Higher Education Personnel (CAPES), and the Ministry of Health (Brazil).

### Author Declarations

This study was approved by the Research Ethics Committee of the Universidade Federal de Santa Catarina

